# Detectable serum SARS-CoV-2 viral load (RNAaemia) is closely associated with drastically elevated interleukin 6 (IL-6) level in critically ill COVID-19 patients

**DOI:** 10.1101/2020.02.29.20029520

**Authors:** Xiaohua Chen, Binghong Zhao, Yueming Qu, Yurou Chen, Jie Xiong, Yong Feng, Dong Men, Qianchuan Huang, Ying Liu, Bo Yang, Jinya Ding, Feng Li

## Abstract

**Background:** Although the SARS-CoV-2 viral load detection of respiratory specimen has been widely used for novel coronavirus disease (COVID-19) diagnosis, it is undeniable that serum SARS-CoV-2 nucleic acid (RNAaemia) could be detected in a fraction of the COVID-19 patients. However, it is not clear that if the incidence of RNAaemia could be correlated with the occurrence of cytokine storm or with the specific class of patients.

**Methods:** This study enrolled 48 patients with COVID-19 admitted to the General Hospital of Central Theater Command, PLA, a designated hospital in Wuhan, China. The patients were divided into three groups according to the Diagnosis and Treatment of New Coronavirus Pneumonia (version 6) published by the National Health Commission of China. The clinical and laboratory data were collected. The serum viral load detection and serum IL-6 levels were determined. Except for routine statistical analysis, Generalized Linear Models (GLMs) analysis was used to establish a patient status prediction model based on real-time RT-PCR Ct value.

**Findings:** The Result showed that cases with RNAaemia were exclusively confirmed in critically ill patients group and appeared to reflect the illness severity. Further more, the inflammatory cytokine IL-6 levels were significantly elevated in critically ill patients, which is almost 10-folds higher than those in other patients. More importantly, the extremely high IL-6 level was closely correlated with the incidence of RNAaemia (R=0.902) and the vital signs of COVID-19 patients (R= −0.682).

**Interpretation:** Serum SARS-CoV-2 viral load (RNAaemia) is strongly associated with cytokine storm and can be used to predict the poor prognosis of COVID-19 patients. Moreover, our results strongly suggest that cytokine IL-6 should be considered as a therapeutic target in critically ill patients with excessive inflammatory response.

## Introduction

The novel coronavirus disease (COVID-19), which is caused by SARS-CoV-2 virus infection, has been called as a public health emergency of international concern by the World Health Organization (WHO)^1-3^. The disease spread rapidly from Wuhan to other areas. Compared with two other WHO blueprint priority coronaviruses, SARS-CoV and MERS-CoV, SARS-CoV-2 is lethal^4^. As of February 25, 2020, SARS-CoV-2 has led to 2663 deaths in China base on the report from National Health Commission of China.

SARS-CoV-2 and SARS-CoV likely use the same angiotensin-converting enzyme 2 (ACE2) as the entry receptor^5-7^, but the clinical manifestations of two diseases were different. COVID-19 patients may develop mild or severe symptoms after infection. The mild patients show symptoms of fever, nonproductive cough, dyspnea, myalgia, fatigue, radiographic evidence of pneumonia; most of them appear to have a good prognosis. In contrast, some patients may develop severe pneumonia, acute respiratory distress syndrome (ARDS) or multiple organ failure^8-11^. Importantly, in most moribund patients, SARS-CoV-2 infection is also associated with an inflammatory cytokine storm^4, 12^, which is mainly characterized by elevated plasma concentrations of interleukins 6 (IL-6). Indeed, several recent COVID-19 clinical studies all indicated that interleukins 6 levels is higher in severe group than those of mild group^12-15^, suggesting that IL-6 might be used as a biomarker for severity evaluation. However, how the IL-6 level is quantitatively correlated with moribund patients is still unknown.

Reverse-transcriptase–polymerase-chain-reaction assay (qPCR) with primers and probes targeting the N and ORF1ab genes of SARS-CoV-2 from throat swab samples have been widely used for diagnosis of COVID-19 patients. Recent investigation showed that the viral load that was determined in the asymptomatic patient was similar to that in the symptomatic patients^16^, implying that the viral load in respiratory specimens may not be able to objectively reflect the disease severity. Serum SARS-CoV-2 viral RNA (termed as RNAaemia by the authors in an recent study) was detected in 15% of the COVID-19 patients ^9^, but the relevant characterizations is lacking. In particular, it is not clear if serum SARS-CoV-2 viral loads could be considered as a prognostic marker, especially for the severe or moribund patients, is not clear.

In this study, we systematically quantify the serum SARS-CoV-2 viral load (RNAaemia) in various patients groups, and characterized the relationship between serum viral loads, interleukins 6 level and disease severity.

## Method

### Data collection

The study was approved by the Ethics Committee of General Hospital of Central Theater Command. Oral consent was obtained from patients. The 48 COVID-19 patients enrolled were confirmed by RT-PCR using throat-swab specimens collected at General Hospital of Central Theater Command between February 01 and February 19, 2020. Their medical records were obtained, including epidemiological, demographic, clinical manifestation, radiological characteristics, laboratory data, and outcome data were obtained. All data were checked by a team of trained physicians.

### Laboratory examination

Serum samples or throat-swabs were collected from all patients and RNAs were extracted. Real-time reverse transcription polymerase chain reaction assay (RT-PCR) was used to determine the viral loads by using a SARS-CoV-2 nucleic acid detection kit (DAAN GENE Ltd., Guangzhou, China, Cat# DA0930-DA0932). Two target genes, the open reading frame1ab (ORF1ab) and nucleocapsidprotein (N),were simultaneously amplified and tested. A cycle threshold value (Ct-value) fewer than 40 was defined as a positive test result, and a Ct-value of 40 or more was defined as a negative outcome according to the manufacturer’s protocol. Specimens, including sputum or alveolar lavatory fluid, blood, urine, and feces, were cultured to identify pathogenic bacteria or fungi that may be associated with the SARS-CoV-2 infection. Inflammatory cytokine IL-6 levels were detected with kit from Roche Ltd (Mannheim, Germany, Cat# 05109442190).

### Statistical Analysis

The categorization of COVID-19 patients were according to the Diagnosis and Treatment of New Coronavirus Pneumonia (Version 6) published by the National Health Commission of China. Classification variables were described as frequency rates or percentages, and significance is detected by chi square or Fisher’s exact test. The quantized variables of parameters are expressed as mean ± standard deviation, and the significance is tested by t-test. SPSS statistical software (Macintosh version 26.0, IBM, Armonk, NY, USA) and R package are used for statistical analysis.

Generalized linear models (GLMs) were used to analyze the degree of patient severity. We first fitted GLMs regarding different response variables with all explanatory factors and their interactions, and then selected the optimum models by applying a stepwise backward selection method via an Akaike information criterion (AIC) index. For the patient’s severity data, four degrees, mild (1), severe (2), critical (3) and dead (4) have been included in the GLM, taking a Gaussian family distribution into account.

## Result

### Characteristics of enrolled COVID-19 patients in this study

A total of 48 patients were enrolled in this study, which are all laboratory confirmed cases. According to the Guidelines of the Diagnosis and Treatment of New Coronavirus Pneumonia (version 6) published by the National Health Commission of China, the enrolled COVID-19 patients were categorized into three groups: 21 mild cases (43.7%), 10 severe cases (20.8%) and 17 critically ill cases (35.4%). Three patients in the critical group died after disease onset. The required conditions must be met for mild patients was the positive SARS-CoV-2 RNA RT-PCR result, because the mild patients might be asymptomatic ^8^, fever or other respiratory symptoms and the typical CT image abnormities of viral pneumonia are optional. Severe patients additionally met at least one of the following conditions: (1) Shortness of breath, RR≥30 times/min, (2) Oxygen saturation (Resting state) ≤93%, or (3) PaO2/FiO2 ≤300mmHg. Enrollment of critically ill patients need to met at least one of the extra following conditions: (1) Respiratory failure that need to receive mechanical ventilation; (2) Shock and (3) Multiple organ failure that need to be transferred to the intensive care unit (ICU).

As shown in **Table 1**, the enrolled COVID-19 patients consisted of 31 males (77.1%) and 17 females (22.9%). The ages of the critically ill patient group (79.6±12.6 years) were older than that of both sever group (63.9±15.2 years) and mild group (45.8±14.2 years), displaying an age-dependent severity. It is noticeable that several underlying diseases were implicated in the COVID-19 patients, the high-risk factors including diabetes (12 [25%]), hypertension (23 [49.7%]) and heart disease (8 [16.7%]). Mixed fungal infection was found in 27.1% patients and the bacterial appear to be uncommon (2.1%).

**Table 1.** Demographics and baseline characteristics of patients infected with SARS-CoV-2.

| Baseline variables | All patients | Mild patients | Severe patients | Critically ill patients | P-value |
| --- | --- | --- | --- | --- | --- |
|  | (N=48) | (N=21) | (N=10) | (N=17) |  |
| Characteristics |  |  |  |  |  |
| Age (year) | 64.6±18.1 | 52.8±14.2 | 63.9±15.2 | 79.6±12.6 | 0.124 |
| Gender (%) |  |  |  |  | <0.001 |
| Men | 37 (77.1) | 13 (61.9) | 9 (90) | 15 (88.2) |  |
| Women | 11 (22.9) | 8 (38.1) | 1 (10) | 2 (11.8) |  |
| Huanan seafood market exposure (%) | 1 (2.1) | 0 (0) | 0 (0) | 1 (5.9) | 0.002 |
| Underlying diseases (%) |  |  |  |  |  |
| Diabetes | 12(25) | 4(19.0) | 1(10) | 7(41.2) | <0.05 |
| Hypertension | 23(49.7) | 6(28.6) | 5(50) | 12 (70.6) | <0.001 |
| Pulmonary disease | 2(4.2) | 1(4.8) | 0(0) | 1(5.9) | 0.043 |
| Hepatic disease | 4(8.3) | 3(14.3) | 0(0) | 1(5.9) | <0.001 |
| Heart disease | 8(16.7) | 2(9.5) | 1(10) | 5(29.0) | <0.001 |
| Cerebral disease | 6(12.5) | 2(9.5) | 0(0) | 4(23.5) | <0.001 |
| Thyroid disease | 4(8.3) | 2(9.5) | 0(0) | 2(11.8) | <0.001 |
| Malignancy | 6(12.5) | 1(4.8) | 2(20) | 3(17.6) | 0.005 |
| Co-infection (%) |  |  |  |  | 0.006 |
| Fungi | 13(27.1) | 1(4.8) | 6(60) | 6(35.3) | <0.001 |
| Bacteria | 1(2.1) | 0(0) | 0(0) | 1(5.9) | 0.004 |

### The serum SARS-CoV-2 nucleic acid was only detectable in critically ill patients

We examined the SARS-CoV-2 viral load in patients serum by real time RT-PCR, as shown in **Figure 1A and B**, 5 out of 48 (10.4%) were confirmed as positive (Ct-value less than 40), which was similar to the previous study^9^. Interestingly, although all the patients were throat-swab specimens test positive, none of the serum sample from mild or severe group showed positive outcome; In contrast, all of the five positive results were from critically ill patients, and two of them died after the onset of the COVID-19 (**Figure 1C and D**). Except for routine statistical analysis, Generalized Linear Models (GLMs) analysis was used to establish an patient status prediction model based on real-time RT-PCR Ct value and their distribution in various patients groups. As shown in **Figure 2**, all the dots that stood for the ORF1ab-qPCR positive patients aggregated and located in the area represent the high probability of death occurs, indicating that RNAaemia is tightly associated with severity.

**Figure 1:**
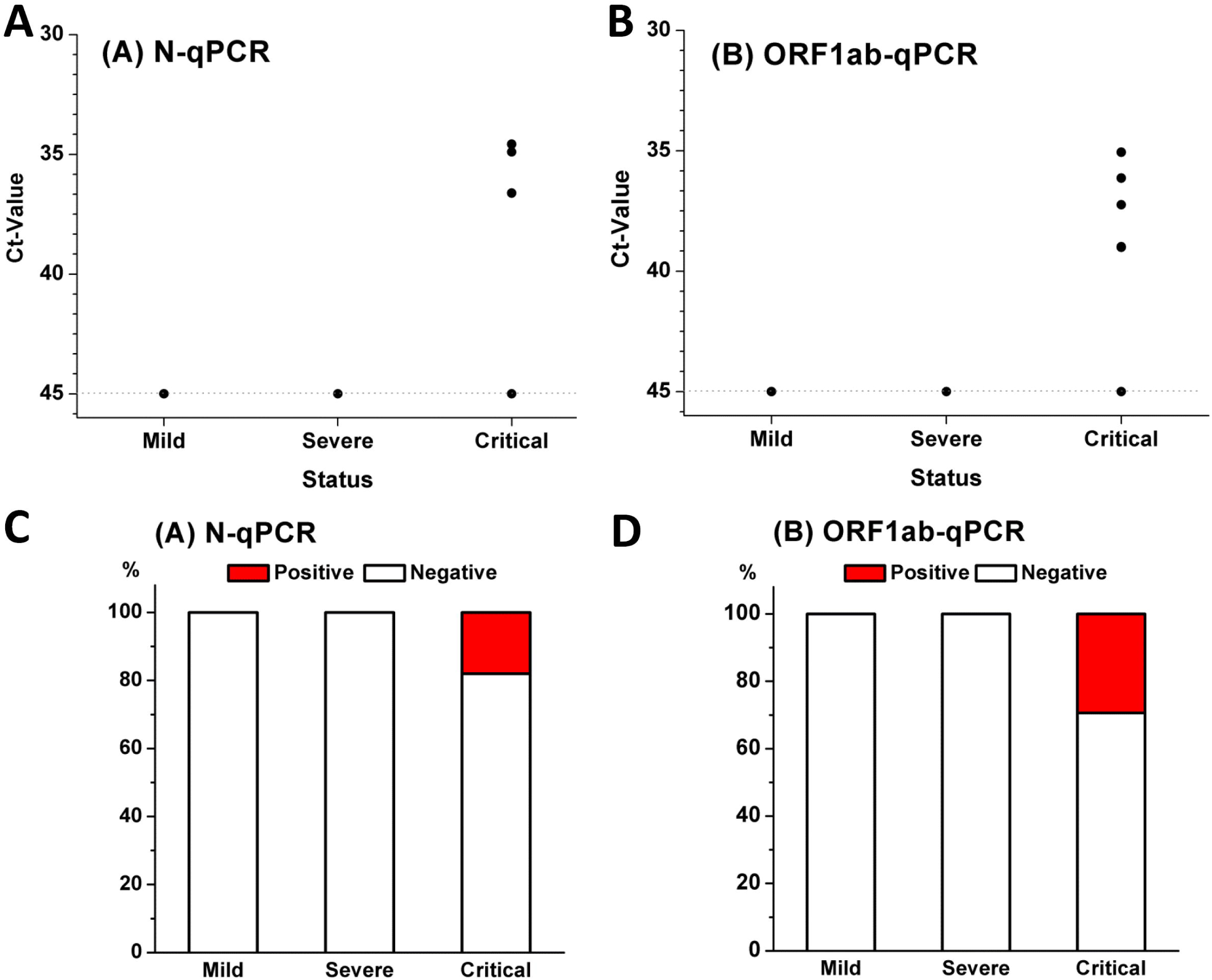
Serum SARS-Cov-2 nucleic acid was exclusively detected in critically ill patients. The scattergram showed the distribution of positive (Ct) values of nucleocapsidprotein (N) (**A**) or (**B**) open reading frame1ab (ORF1ab) in various patients groups. The histogram indicates the ratio of cases with positive (Ct) values of (**C**) nucleocapsidprotein (N) or (**D**) open reading frame1ab (ORF1ab) in each of the patients groups.

**Figure 2:**
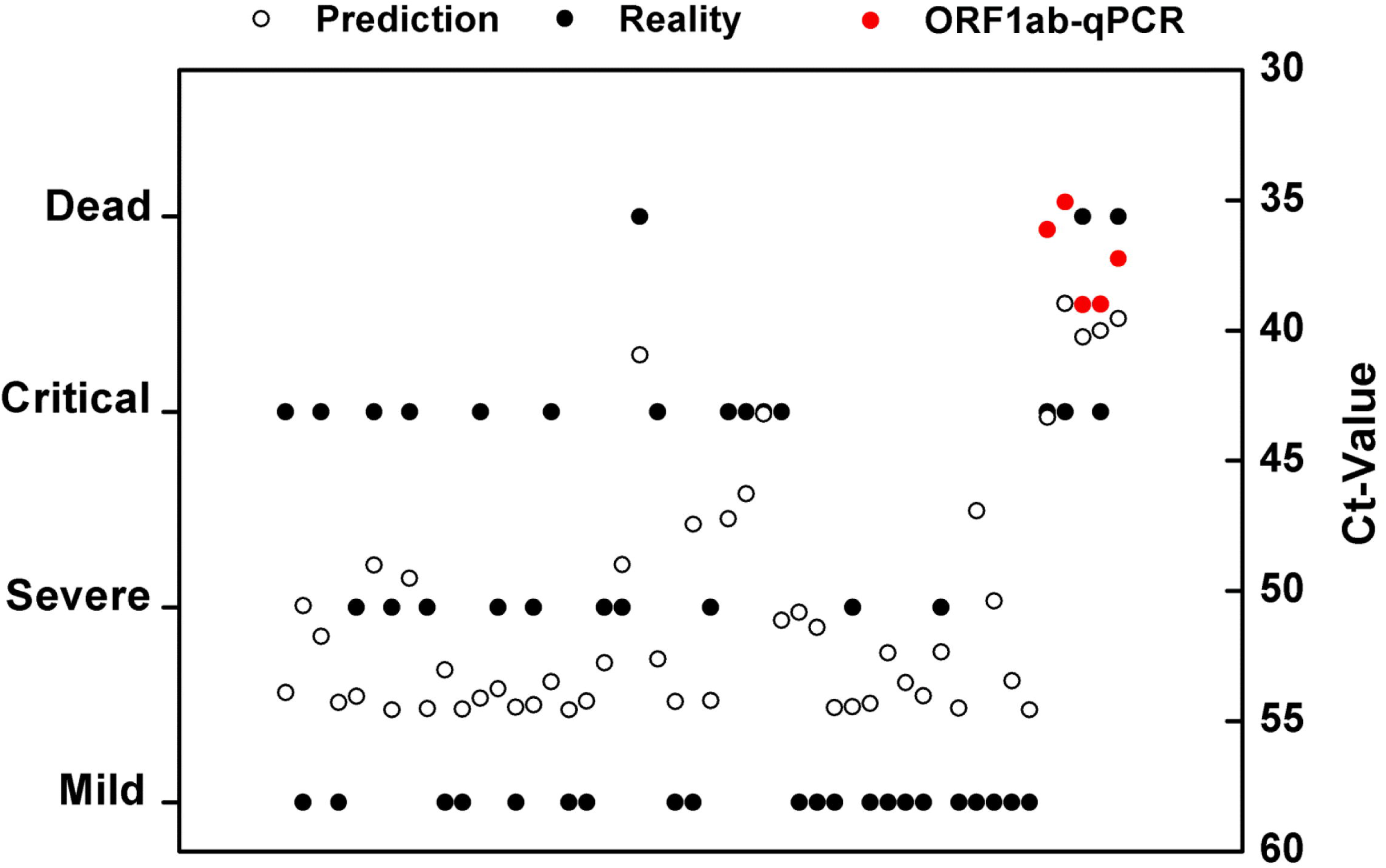
Generalized Linear Models (GLMs) analysis was used to establish an patient status prediction model based on real-time RT-PCR Ct value and their distribution in various patients groups. Mild, mild patients; Severe, severe patients; Critical, Critically ill patients;

Since there is no direct evidence that SARS-CoV-2 could infect the blood cells, and serum virus load appear to reflect the severity, we assumed that the serum virus RNA probably was derived from the damaged organs and ruptured vessel, which was caused by aberrant and excessive immune responses. Because inflammatory cytokine storm is frequently occurred in critically ill COVID-19 patients, we then asked that if there any laboratory parameters could be associated RNAaemia and contribute to the COVID-19 severity?

### Sharply increased IL-6 level was strongly associated with the COVID-19 severity

As shown in **Table 2**, the absolute counts of lymphocytes in the peripheral blood of the severe patients was lower, and became even lower in critically ill patients, which is consistent with recent report^17, 18^, while the absolute counts of neutrophils were higher in critically ill patients. Procalcitonin (PCT) value was higher in critically ill patients compared with another two groups, which was similar to other clinical observation^19^, indicating an obvious elevated inflammatory response in those patients. Remarkably, sharply increased IL-6 level was observed in critically ill patients, which was almost 10-folds higher than that of severe patients, moreover, all of the death cases exhibited extremely high IL-6 value(**Table S1**), suggesting that IL-6 might be an important biomarker of poor prognosis in COVID-19 patients. The extremely high level of IL-6 is a hallmark and an important driving force of cytokine storm^20^, which may cause multiple organs functional disability in critically ill patients^4^. Consistently, the parameters stand for the organ dysfunction, including TnT (troponin T), AST (aspartate transaminase), ALT (aspartate transaminase), CRE (serum creatinine), and BUN (blood urea nitrogen), all appear to be higher in critically ill patients compared with another two groups.

**Table 2.** Comparison of laboratory parameters in mild, severe and critically ill COVID-19 patients.

| Baseline variables | All patients<br>(N=48) | Mild patients<br>(N=21) | Severe patients<br>(N=10) | Critically ill<br>patients<br>(N=17) | P-value |
| --- | --- | --- | --- | --- | --- |
| White blood<br>cell($\times 10^9/L$) | 6.7±3.8 | 5.5±2.2 | 5.1±3.0 | 9.1±4.4 | <0.05 |
| Neutrophil($\times 10^9/L$) | 3.8(2.7-6.6) | 3.4(2.8-4.3) | 2.9(2.0-3.78) | 7.1(5.3-9.2) | <0.05 |
| Lymphocyte($\times 10^9/L$) | 0.9(0.6-1.3) | 1.2(1.0-1.4) | 0.8(0.7-0.9) | 0.4(0.3-0.6) | <0.05 |
| IL-6(pg/ml) | 18.1(4.5-49) | 10.4(3.8-31.0) | 5.8(3.1-16.9) | 64(25.6-111.9) | <0.001 |
| PCT(ng/ml) | 0.06(0.04-0.13) | 0.04(0.03-0.06) | 0.04(0.04-0.06) | 0.2(0.1-0.6) | <0.05 |
| TnT(ng/ml) | 0.01(0.008-0.03) | 0.008(0.005-0.01) | 0.009(0.007-0.01) | 0.03(0.02-0.06) | <0.001 |
| ALT(U/L) | 31(16.8-44.3) | 36.0(15.0-48.0) | 20.5(16.5-29.3) | 32.0(19.0-50.0) | <0.05 |
| AST(U/L) | 34.0(25.0-43.5) | 34.0(25.0-43.0) | 27.5(24.5-29.5) | 42.0(25.0-60.0) | <0.05 |
| CRE(umol/L) | 64.5(55.0-77.0) | 63.0(50.0-71.0) | 56.0(52.0-73.8) | 77.0(64.0-101.0) | <0.05 |
| BUN(mmol/L) | 5.6(3.8-7.9) | 4.0(3.5-5.1) | 4.2(2.6-7.8) | 9.3(7.5-13.7) | <0.05 |

Abbreviations: PCT, procalcitonin; IL-6, interleukin-6; TnT, troponin T; ALT, alanine aminotransferase; AST, aspartate aminotransferase; CRE, serum creatinine; BUN, blood urea nitrogen.

### RNAaemia is closely associated with IL-6 level in critically ill COVID-19 patients

RNAaemia has been proposed in recent study as a COVID-19 related phenomenon. Our data strongly suggest that both RNAaemia and high IL-6 level were exclusively observed in critically ill patients, which prompt us to further study the relation between them. As shown in **Figure 3A**, patients with RNAaemia exhibited much higher IL-6 level compared with other patients. We then checked the IL-6 value in each of the RNAaemia patients, strikingly, all of their IL-6 value were more than 100 (**Table S1**). In order to further confirm the relation between them, we firstly analyze the IL-6 value in the critically ill patients, notably, the mortality appeared to be correlated with IL-6 value ≥100 because all of the death in this study were belong to this group (**Table S1**), we therefore define IL-6 value ≥100 as high and the rest of cases as low, the IL-6 high patients accounted for 35.3% in critically ill group (**Figure 3B**). As shown in **Table 3**, the incidence of RNAaemia was closely correlated with IL-6 high in critically ill patients (R=0.902). These data clearly suggested that RNAaemia was related with poor prognosis, indeed, all the RNAaemia patients were at higher risk of multiple organs impairment compared to those patients without RNAaemia (**Figure 4A-C**).

**Table 3.** The correlation analysis of RNAaemia incidence or vital signs and serum IL-6 level in 48 COVID-19 patients.

| Baseline variables | All patients<br>(N=48) | IL-6 (< 100 pg/ml)<br>(N=42) | IL-6 (> 100pg/ml )<br>(N=6) | R | P-value |
| --- | --- | --- | --- | --- | --- |
| RNAemia |  |  |  | 0.902 | <0.001 |
| negative | 43 | 42 (100) | 1 (16.7) |  |  |
| positive | 5 | 0 (0) | 5 (83.3) |  |  |
| Vital signs |  |  |  | -0.683 | 0.001 |
| death | 3 | 0(0) | 3(50) |  |  |
| alive | 45 | 42(100) | 3(50) |  |  |

**Figure 3:**
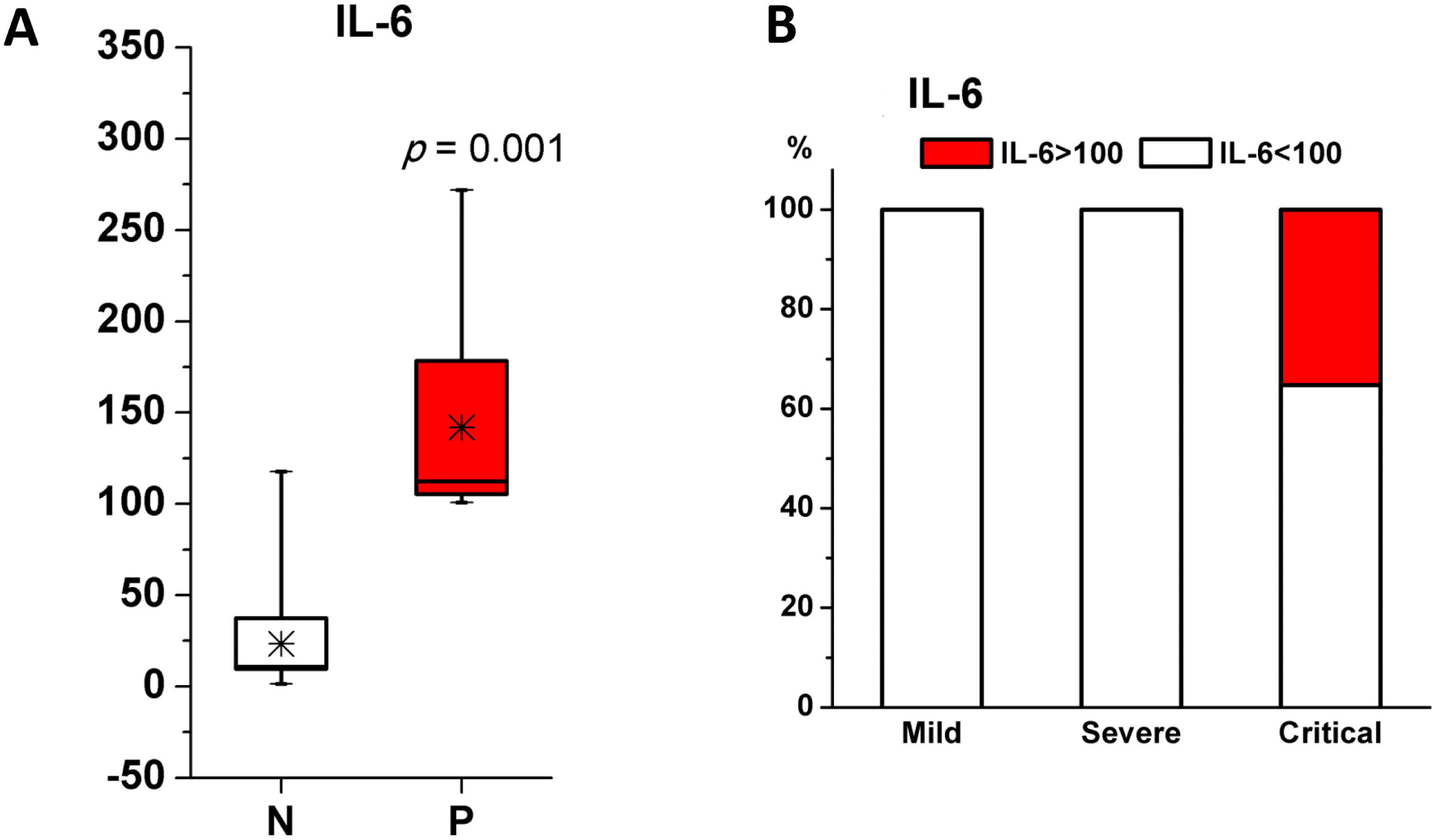
(**A**) The average IL-6 value in cases with (P) or without (N) RNAaemia; (**B**) The ratio of patients with high expression level of IL-6 (IL≥100) is each of the group.

**Figure 4:**
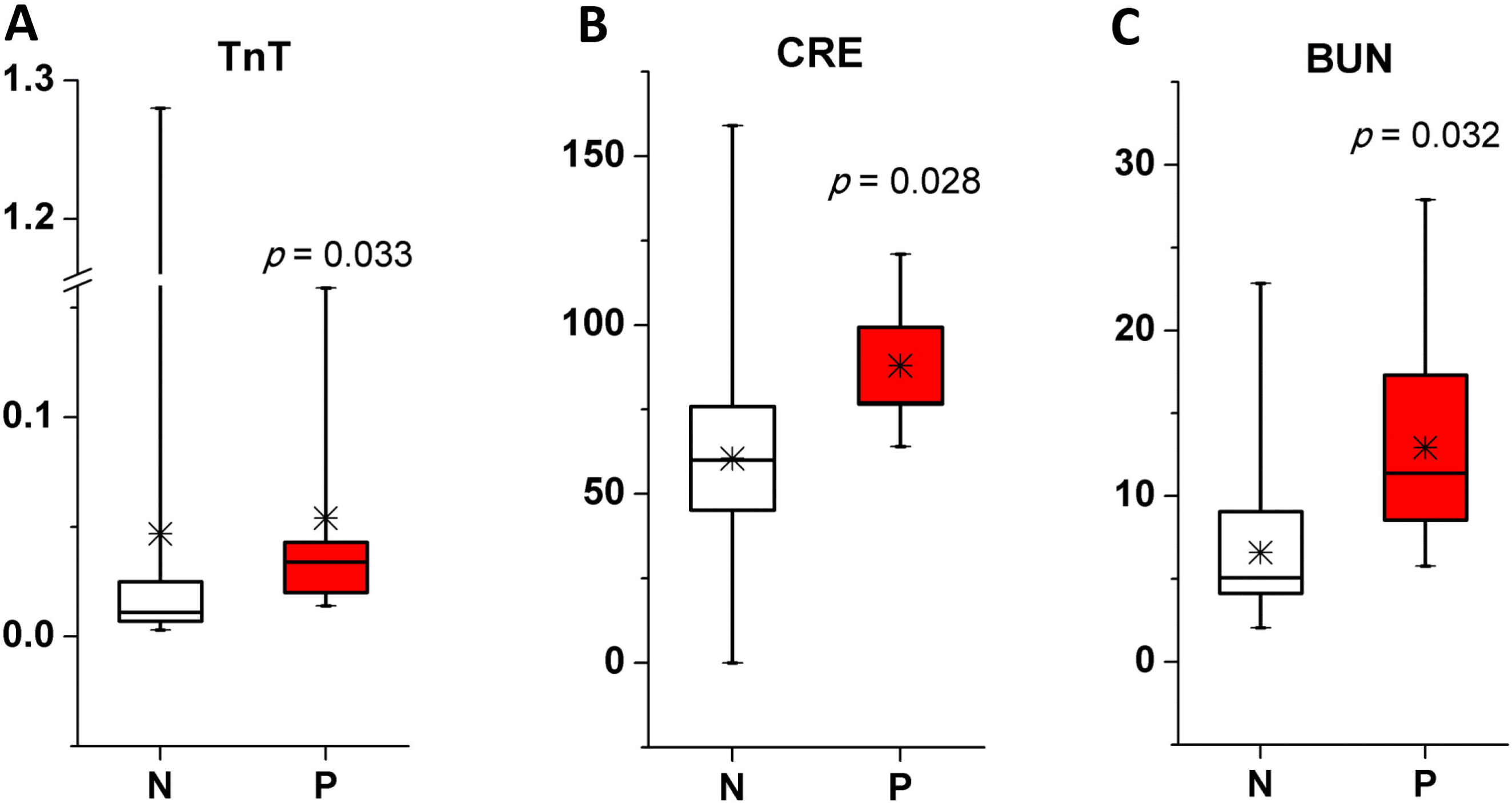
The average value of TnT, CRE and BUN in cases with (P) or without (N) RNAaemia. TnT, troponin T; CRE, serum creatinine; BUN, blood urea nitrogen. Mild, mild patients;

## Discussion

Although the detection of throat-swab rather than serum SARS-CoV-2 viral load is widely used for the COVID-19 diagnosis, an undeniable fact is that serum SARS-CoV-2 nucleic acid (RNAaemia) is detectable in part of the patients, however, in what scenario it become detectable and whether the incidence of RNAaemia could be correlated with specific type of patients is not very clear. In this study, we investigated the distribution of serum SARS-CoV-2 nucleic acid positive cases in various patients groups, and the result showed that those cases all were confirmed only in critically ill patients group. Moreover, the laboratory data analysis strongly suggest that inflammatory cytokine IL-6 level was significantly elevated in critically ill patients. More importantly, the extremely high IL-6 level was closely correlated with the incidence of RNAaemia and the mortality. Our work may provide clues for developing new COVID-19 diagnostic strategy and therapeutic targets.

Although recent studies have shown that IL-6 level increased in severe patients^14^, its level in COVID-19 critically ill patients is still unknown. IL-6 is one of the major pro-inflammatory factors that contribute to the formation of cytokine storm, which largely enhance the vascular permeability and impair the organs function. This observation might help to explain that why serum SARS-CoV-2 viral RNA was only able to be detected in patients with extremely high level of IL-6. We still could not rule out the possibility that SARS-CoV-2 virus amount grow explosively in a short period, which in turn induced the massive cytokine storm that characterized by elevated level of cytokines such as IL-6. We have to point out that due to the shortage of detection kit, the results of other cytokines could not be obtained and those investigations should be warranted in future study.

Our research indicated that serum SARS-CoV-2 viral load and IL-6 level could serve as an indicator of poor prognosis. Since the mortality of COVID-19 critically ill patients is considerable, the host-directed therapies should be an option. If IL-6 could become a therapeutic target for treatment of critically ill patient? Notably, the IL-6 monoclonal antibody-directed COVID-19 therapy has been used in clinical trial in China (No.ChiCTR2000029765). Our data strongly support this notion and efficacy of IL-6 monoclonal antibody directed therapy remains to be fully evaluated.

## Data Availability

All data referred to the manuscript are available to readers once this submission is posted online.

## Acknowledgment

This work was supported by grants from the National Natural Science Foundation of China (91859206); Medical Science Advancement Program (Basic Medical Science) of Wuhan University (TFJC2018003); Wuhan University Funding (2042019kf1018). We thank Louise T. Chow (University of Alabama at Birmingham) for suggestions and critical reading of this manuscript.

## Contributors

FL and JD conceived and designed the study. FL, XC, YF and DM contributed to the literature search. XC, BZ, QH,YL, BY and JD contributed to data collection. XC,YQ, YC and FL contributed to data analysis. FL and XC contributed to data interpretation. YQ and XC contributed to the figures. FL,XC and JX contributed to writing of the report.

